# A study protocol for an international registry observational study evaluating clinical outcomes of transcatheter versus standard surgical mitral valve operation for secondary mitral regurgitation: the TEERMISO study

**DOI:** 10.1101/2024.11.01.24316548

**Authors:** Francesco Nappi, Sanjeet Singh Avtaar Singh, Antonio Salsano, Cristiano Spadaccio, Yasushige Shingu, Satoru Wakasa, Antonio Fiore

## Abstract

**Background:** Secondary mitral regurgitation (SMR) is a condition affecting the left ventricle (LV) rather than the mitral valve (MV). If the mitral valve (MV) remains structurally unchanged, enlargement of the left ventricle (LV) or impairment of the papillary muscles (PM) can occur. Several mechanical interventions are available to dictate the resolution of MR. However, there is a lack of robust data to compare mitral valve replacement, mitral valve repair (including subvalvular repair), and transcatheter mitral valve procedures (TMVp). This study aims to compare the effectiveness and clinical outcomes of TMVp using the edge-to-edge mitral valve repair (TEER) technique and standard surgical mitral valve procedures (S-SMVp) in patients with SMR.

**Methods and analysis:** Five cardiac surgery centres from four European countries and Japan have collaborated to create a multicentre observational registry (TEERMISO). The registry will enrol consecutive patients who underwent mechanical intervention for SMR between January 2007 and December 2023. The investigators assessed the difference between replacement and repair for both the standard surgical approach and the transcatheter procedure. The main clinical outcome will be the degree of LV remodelling as assessed by the Left Ventricular End-Diastolic Volume Index at 10 years. The study will measure several secondary endpoints, including all-cause mortality as the primary endpoint, followed by functional status, hospitalisation, neurocognition, physiological measures (echocardiographic assessment), adverse events and reoperation.

**Ethics and dissemination:** Ethics approval was obtained in Montpellier University Hospital on 24 May 2022 (Institutional Review Board (IRB) Approval Number: IRB-MTP_2022_05_202201143). The results of the main study and each sub-analysis will be submitted for publication in a peer-reviewed journal. ClinicalTrials.gov ID: NCT05090540; IRB ID: 202201143. (Supplementary material)

**Strengths and limitations of this study:**

- This study will be conducted as a large international registry concerning interventions to correct secondary mitral regurgitation; it will provide clinicians important information about transcatheter and surgical techniques in the specific field of the secondary mitral regurgitation.
- The primary outcome will offer opportunity to better predict left ventricular remodeling after the procedures.
- The secondary outcomes of this study will offer opportunity to provide important information abut survival
- The retrospective nature of the study is a limitation to the study design.

## 1. Introduction

Secondary mitral regurgitation (SMR) occurs when the left ventricle (LV) becomes enlarged or the papillary muscles are impaired, without any structural changes to the mitral valve (MV). Guideline-directed medical therapy (GDMT), cardiac resynchronization, and coronary revascularization are primary treatments for heart failure (HF) patients with SMR to improve the substrate LV dysfunction. As a disease of the LV rather than the MV, the impact of therapeutic measures to improve SMR on subsequent clinical outcomes is debated. [1–3] There is no definitive evidence to support surgery for the treatment of SMR. Although Restrictive mitral annuloplasty (RMA) is the most commonly used conservative technique for the standard surgical mitral valve procedure (S-SMVp), observational studies have shown that it is effective in reducing SMR and improving symptoms. However, it is uncertain whether these results are sustainable or whether it improves mortality compared with GDMT. [4–7] It is important to further investigate the long-term effects of RMA on patient outcomes.

In addition, there are different surgical techniques for the treatment of MV disease. While a randomized controlled trial found that SMV-r was associated with comparable survival rates, it also showed a higher risk of recurrent MR and cardiovascular readmission compared to chordal-sparing mitral valve replacement. However, the choice of procedure still largely depends on the experience and practice of the center and operator. [1–3] In light of the limited evidence of clinical benefit, surgeons may be reluctant to employ two-stage mitral valve repair, which combines subvalvular papillary muscle repair (SPM-r) with RMA, as an alternative technique for handling the MV. This is based on observational studies [8,9] and a small number of randomized clinical trials (RCT). [10,11]

Recently, two randomized controlled trials have been conducted: the Multicenter Study of Percutaneous Mitral Valve Repair MitraClip Device in Patients With Severe Secondary Mitral Regurgitation (MITRA-FR) and the Cardiovascular Outcomes Assessment of the MitraClip Percutaneous Therapy for Heart Failure Patients With Functional Mitral Regurgitation (COAPT). The effectiveness of transcatheter mitral valve procedure (TMVp) using the edge to edge MitraClip system (Abbott, Abbott Park, Illinois) in combination with GDMT was evaluated and compared with GDMT alone. [12–15] In the MITRA-FR RCT, the addition of percutaneous repair to medical therapy did not significantly reduce the risk of death or HF hospitalisation compared with medical therapy alone at 1 [12] and 2 years. [13] However, in the COAPT RCT, patients with heart failure and moderate-to-severe or severe secondary mitral regurgitation who remained symptomatic despite guideline-directed medical therapy, transcatheter edge-to-edge mitral valve repair (TEER) was safe. It led to a lower rate of hospitalisation for heart failure and all-cause mortality at 2 [13,14] and 5 years of follow-up than medical therapy alone. [13,15] COAPT is the first randomized controlled trial to suggest a survival benefit from correcting SMR through TEER.

Limited data exist comparing TEER with conventional surgery for treating SMR, including the surgical procedure aimed at correcting papillary muscle dislocation. One study used propensity score matching, but it included the surgical technique of RMA [16], while the other did not use propensity score matching and focused on the edge-to-edge surgical approach [17] and mitral valve replacement. [18] Our hypothesis is that TEER and S-SMVp may result in different survival rates, functional outcomes, and echocardiographic results for the treatment of SMR. The multicenter registry of transcatheter Versus Standard Surgical Mitral Valve Operation for Secondary Mitral Regurgitation (TEERMISO) aims to evaluate the clinical outcomes of patients undergoing TEER using the MitraClip system compared to those undergoing S-SMVp using different procedures for mitral valve repair and replacement for the treatment of SMR. The study compare the long-term clinical outcomes of two propensity score-matched cohorts using LASSO regression on a large population cohort.

## 2. Methods and analysis

The TEERMISO is an observational registry that includes patients who underwent mechanical intervention for SMR at five cardiac surgery centers located in two European countries (France and Italy) and Japan (**Table 1**). During the study period, we will review data on patients who have been treated. The purpose is to provide more data for future clinical research on this topic. Data on consecutive patients with SMR will be gathered in a Microsoft Access datasheet (Redmond, Washington, USA). The datasheet will include prespecified baseline, operative, and outcome variables. The study started in 2007 and patients will be enrolled until 2024, depending on the results of consecutive interim analyses. Permission to conduct this study will be requested from the institutional review board or local ethical committee in accordance with local legislation.

**Table 1.**
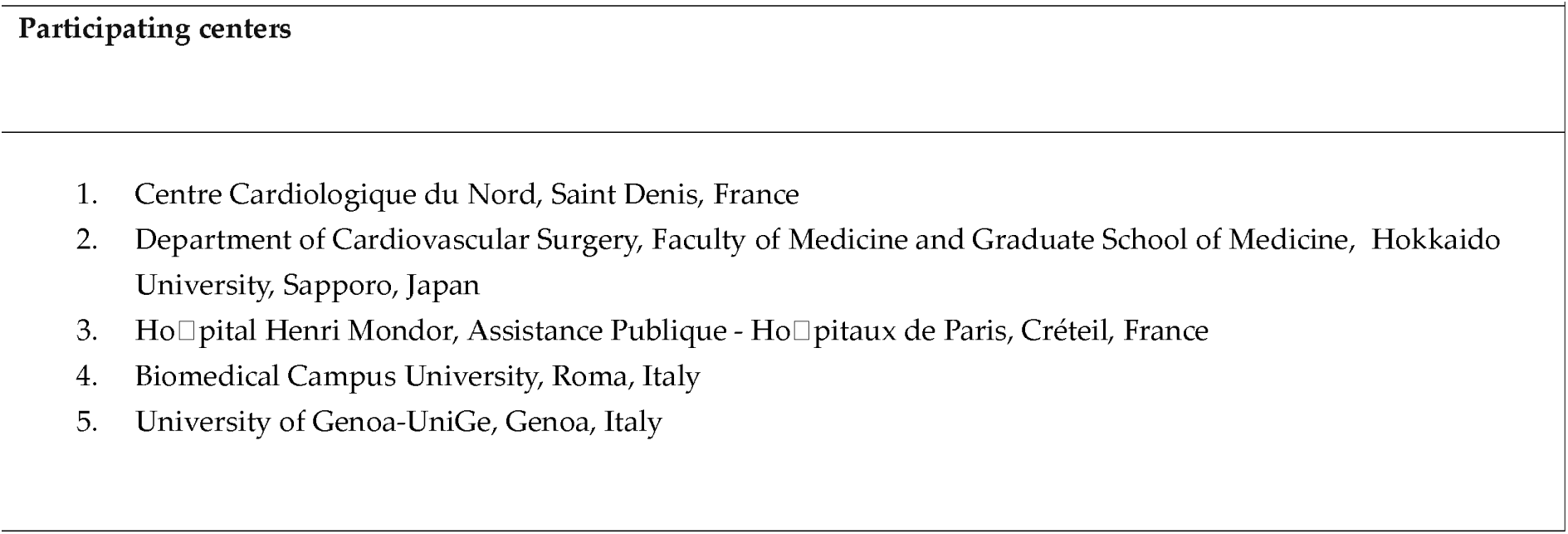
Participating centers.

### 2.1. TEERMISO Study Patient Entry Criteria

#### Characterization of patient populations

The trial will include patients who have moderate and severe mitral regurgitation, with or without the need for concomitant coronary artery bypass surgery (CABG) or percutaneous coronary intervention (PCI), and who have undergone either surgery or TMVp using TEER. Patients of any gender, race or ethnicity are eligible for inclusion in the study

#### Inclusion criteria

- Age 18 years or older
- Chronic severe ischemic and non-ischemic mitral regurgitation (ERO ≥ 0.4 cm echocardiogram) with tethering as a major mechanism.
- Symptomatic secondary mitral regurgitation (3+ or 4+ by echocardiographic laboratory assessment) due to cardiomyopathy of either ischaemic or non-ischaemic etiology.
- The subject has been adequately managed according to applicable guidelines, including for coronary artery disease, LV dysfunction, mitral regurgitation and heart failure.
- Managed with standard surgical mitral valve procedure or transcatheter mitral valve procedure
- Coronary artery disease in patients who have undergone a S-SMVp, with or without the need for coronary revascularization.
- The cardiac team at each centre will determine which patients to include in the register. This will be based on those who have undergone standard mitral valve surgery or a transcatheter mitral valve repair procedure.
- To participate in the TEERMISO registry, the individual must be capable of signing both the Informed Consent and Release of Medical Information forms.

#### Exclusion Criteria

- Pediatric
- Any echocardiographic evidence of structural (chordal or leaflet) mitral-valve disease
- Papillary muscle rupture
- Infective endocarditis

Based on a survey of the clinical centers involved in this study, it is estimated that the total volume of patients to be evaluated will be 630 patients screened from the databases of the five centers involved in enrolment ***(supplementary material***). Patients will be followed up using strategies that may include mailings to the referring physicians of patients at the hospitals for procedure, checks directly at the centre where the patient underwent surgery, and telephone calls to the healthcare facilities where the patients were eventually readmitted in the case of adverse events. The study centres regularly assessed the actual inclusions against the pre-specified targets. The screening protocol, which covered all centres, recorded the number of patients who underwent different procedures and received echocardiographic and clinical follow-up.

The database contains information on the participation of women and minorities in clinical studies. This is important for scientific, ethical, and social reasons, as well as for the generalizability of study results. TEERMISO is dedicated to achieving scientific results while ensuring balanced recruitment of patients regardless of gender or ethnicity. The TEERMISO registry has recruited at least 30% women and 25% minorities. To ensure adequate representation of these groups, the recruitment centres implemented two measures: (1) documenting the number of women and minorities selected and enrolled through the screening and exclusion protocols, and (2) monitoring these records by each clinical centre based on annual follow-ups. The included and excluded criteria for patients who underwent mechanical intervention for SMR are shown in **Supplementary Table 1.**

### 2.2. Trial Design and Endpoints

The trial design schematic is reported in **Supplementary** Figure 1.

### 2.3. Therapeutic Interventions

#### Standard Surgical Mitral Valve Procedure (S-SMVp) *(S2 supplementary material)*

##### Mitral Valve Replacement

Mitral valve replacement involves preserving the subvalvular apparatus to prevent left ventricle dilation over time. Surgeons choose the preservation technique, prosthetic valve type, and suture placement based on their preference. In instances of ischaemic cardiomyopathy, CABG is a requisite procedure for revascularisation.

##### Restrictive Mitral Annuloplasty

RMA can be performed using a complete rigid or semi-rigid annuloplasty ring that has been downsized to fit the annulus diameter. As patients who undergo RMA often have coronary artery lesions, a CABG operation may be necessary to ensure favourable remodelling of the left ventricle.

##### Restrictive Mitral Annuloplastie Plus Subvalvular Repair

RMA+SPM-r allows for the approximation or relocation of papillary muscles that are displaced. In patients who have received RMA+SPM-r due to ischemic cardiomyopathy, a CABG operation is required.

##### Leaflet and Chordal Procedures in Functional Mitral Regurgitation

Leaflet and chordal procedures (LCP) are newer repair techniques in functional mitral regurgitation. Results of studies with leaflet augmentation and secondary chordal cutting are promising.

#### Transcatheter Mitral Valve Procedure (TMVp) *(S2-Supplementary material)*

##### Transcatheter Edge to Edge Repair (TEER)

The TEER procedure involves bringing together the edges of the anterior and posterior leaflets of the mitral valve, known as edge-to-edge repair. If one MitraClip device is not enough to reduce mitral regurgitation adequately, a second device may be used to optimize the procedure.

### Endpoint Definitions and Measurement

#### Primary Endpoint

The trial’s primary endpoint is the degree of left ventricular remodeling, which will be assessed by measuring the change in Left Ventricular Ejection Fraction (LVEF) and Left Ventricular End Diastolic Volume Index (LVESVI) at hospitalization and every year after the procedural intervention using transthoracic echocardiography.

#### Secondary Endpoints

The principal secondary endpoint is survival which will be assessed through all-cause mortality. The trial has additional secondary endpoints defined as follows:

#### Perioperative Measures

The following parameters will be measured: operative time, cardiopulmonary bypass time, cross clamp time, blood loss, and transfusions.

##### Cardiopulmonary bypass parameters

Data regarding the duration of myocardial ischemia, cardiopulmonary bypass, and retrograde or antegrade cardiac cardioplegia perfusion will be collected.

##### Blood loss and transfusions

Data on the number of red blood cell units transfused will be collected. A simplified version of the E-CABG perioperative bleeding classification will be adopted, [19] which has been shown to be comparable to the Universal Definition of Perioperative Bleeding [20] in predicting early mortality [21]. Major bleeding is defined as transfusion of more than 4 units of RBC during and after the procedure and/or re-operation due to excessive intrathoracic bleeding. **Table 2**

**Table 2.** Classification for perioperative bleeding in simplified E-CABG.

##### Reoperation for bleeding

A re-opening of the chest for excessive bleeding is a re-operation for bleeding. A reoperation for bleeding refers to any instance where the sternum was left open and subsequent surgery is required to address excessive bleeding. It is important to note that reopening the chest for haemodynamic instability without excessive bleeding and pericardial/pleural puncture or chest tube insertion for retained blood syndrome are not considered reoperations for bleeding.

### Functional Status

#### MACE (Major Adverse Cardiac Events)

MACE is specified as an unweighted composite score. It consists of the following components

`o` Death
`o` Stroke
`o` Mitral valve re-intervention
`o` Worsening heart failure (+1 NYHA Class)
`o` Chronic HF hospitalization

#### New York Heart Association (NYHA) Classification

Functional status is determined using the NYHA classification scale. Full guidelines for the NYHA classification can be found in supplementary material. **Table 3**

**Table 3.**
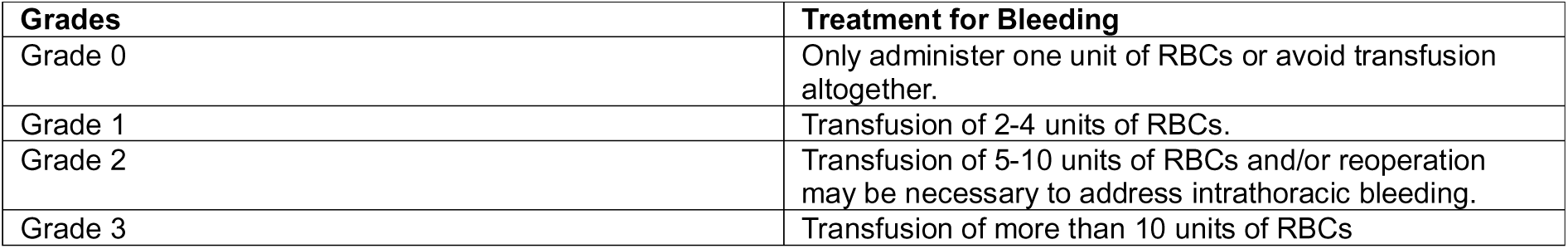
Classification for perioperative bleeding in simplified E-CABG [54].

#### Angina Class

Angina class is assigned according to the Canadian Cardiovascular Society Classification (CCSC). The CCSC guidelines are given in detail in supplementary material. [22] **Table 4**

**Table 4.** New York Heart Association (NYHA) Classification.

| **Class** | **Patient Symptoms** |
| --- | --- |
| Class I (Asymptomatic) | There are no restrictions on physical activity. Normal physical activity should not cause excessive tiredness, heart palpitations, or shortness of breath. |
| Class II (Mild) | The individual experiences a slight limitation in physical activity. They are comfortable at rest, but ordinary physical activity leads to fatigue, palpitation, or dyspnea. |
| Class III (Moderate) | The patient experiences a significant reduction in their ability to engage in physical activity. While they may feel comfortable at rest, any activity beyond the ordinary level leads to fatigue, palpitations, or difficulty breathing. |
| Class IV (Severe) | Any physical activity exacerbates the discomfort. The individual experiences discomfort when engaging in physical activity and displays symptoms of cardiac insufficiency even at rest. |

#### Re-operation

All re-operations and re-operation for mitral regurgitation in particular will be recorded and freedom from re-operation will be analyzed using a time-to-event analysis.

#### Peak VO2

In contrast, functional status will be measured by maximal oxygen uptake (VO2 peak) assessed by cardiopulmonary exercise testing at pre-specified time intervals in patients without a clinical contraindication to exercise testing (e.g. unstable angina or left main disease). All patients will be uniformly prompted to exercise to the point of voluntary exertion by means of a consistent verbal prompting protocol. The addition of Borg’s CR10 RPE (rate of perceived exertion) would be useful in those patients who are unable to reach an RER (respiratory exchange ratio) ≥ 1.0 to complement the level of exertion achieved. For this reason, the RPE is assessed and recorded every 2 minutes for all subjects using the RPE scale from 0 to 10. Scripted patient instructions on the use of the RPE scale will be used for all study related exercise testing, and uniform laminated cards depicting the scale and demonstrating its use will be presented to each patient during testing. Cardiopulmonary exercise testing will be performed in accordance with a uniform protocol and will be interpreted by the cardiologist in charge at each enrolment centre.

### Neurocognition

Neurocognition will be the focus of comparison between treatment groups by assessing cognitive performance using a battery of tests. These tests include the Hopkins Verbal Learning Test, Trail making Tests A and B, MCG Complex Figures, Boston Naming Test, Digit Span, and Digit Symbol Substitution Test Clinical site personnel, trained by experienced neuropsychologists identified at each site of recruiting, will administer neurocognitive testing. Neurologists at each center of enrollment will perform all neurocognitive test scoring.

### Hospitalization

#### Index Hospitalization

We will measure the length of stay for the index hospitalization and break it down by days stayed in the intensive care unit (ICU). Additionally, we will record the discharge location.

#### Readmission

Readmission ratios will be computed for the first 30 days after the procedure and for the duration of follow-up. Hospitalizations will be categorized for all conditions, including cardiovascular and heart failure readmissions. To classify a readmission as heart failure related, at least two of the following signs and conditions of acute decompensated heart failure must be observed:

- Dyspnea felt related to HF
- Administration of vasodilators, intravenous diuretics or inotropes
- Pulmonary capillary wedge pressure (PCWP) or Left ventricular end-diastolic pressure (LVEDP) > 18 mmHg
- On physical examination, rales may indicate the presence of pulmonary edema or pulmonary vascular congestion as seen on X-ray.

The investigator will classify all readmissions, which will then be adjudicated by the cardiologist in charge of hospitalization.

#### Days Spent Alive and Outside of Hospital

The study will compare the total number of days alive and out of the hospital between the treatment groups, as well as the percentage of days out of the hospital in relation to the total days alive post-procedure.

### Physiologic Measures

#### Echocardiographic Tests

`o` Mitral regurgitation will be assessed by measuring the effective regurgitant orifice area (EROA) using both the proximal isovelocity surface area (PISA) and quantitative flow methods to determine its presence and severity.
`o` The mitral valve apparatus and valve area will be evaluated by assessing annular shape and motion, tethering angle and tenting area, papillary muscle location and distance, and calculating the mean trans-mitral stenotic gradient using mitral inflow continuous wave Doppler.
`o` LV dimension, geometry and function will be evaluated by the following parameters: LV dimensions, ejection fraction (biplane Simpson’s rule), LV end-systolic volume index using the biplane volumetric method, LV mass, LV sphericity, radial strain, and twist.
`o` Left atrial size, mitral valve tethering length and area, tethering angle, papillary muscle position and separation will be evaluated echocardiographically.
`o` RV dimension and function will be estimated by the following indices: tricuspid annular plane systolic excursion (TAPSE), peak systolic velocity, diastolic E and A velocities (by tissue Doppler) and fractional area change.
`o` Doppler flow testing will be used to assess intracardiac pressures and hemodynamics, particularly pulmonary artery pressures and pulmonary capillary wedge pressures.
`o` Regional Wall Motion (LV function and viability assessment) will be evaluated at baseline and one year after surgery to assess the extent of baseline viability and the effect of revascularisation, if any.

#### Appropriateness of revascularisation

Preoperatively, coronary territories are identified as either suitable or unsuitable for bypass, and their bypass status will be monitored postoperatively. Identify regions receiving coronary flow as follows: left anterior descending (LAD) proximal, LAD distal, proximal diagonal, distal diagonal, proximal circumflex, distal circumflex, distal dominant circumflex, right posterolateral, and right posterior descending.

### Quality of Life

The study will assess improvement in quality of life (QOL) from baseline using the disease-specific Minnesota Living with Heart Failure (MLHF) score, the disease-specific Duke Activity Status Index (DASI), the Short Form-12 general health status index and the EuroQol 5-D measures of health state preference from an individual and societal perspective. The Minnesota Living with Heart Failure Questionnaire is a tool that measures the physical, psychological, and social eOects of heart failure and its treatment on patients. The DASI questionnaire provides cardiovascular stress across four spheres of adult activity that are linked with oxygen uptake: walking, personal care, household tasks, and sexual function and leisure. The SF-12 examines 8 dimensions of quality of life, including physical activity, social activity, role/physical, body pain, general mental health, role/emotional, vitality, and general health perception. The SF-12 and EuroQoL 5-D are both instruments used to measure health-related quality of life. The EuroQoL 5-D is a standardized questionnaire that provides a simple descriptive profile consisting of 5 dimensions. The EuroQoL 5-D is a standardized questionnaire that provides a simple descriptive profile consisting of 5 dimensions. The instrument assesses five domains: anxiety/depression, pain/discomfort, usual activities, self-care, and mobility. Additionally, it includes a self-assessment of health status.

### Safety

#### Re-operation

All re-operations and re-operation for mitral regurgitation in particular will be recorded and freedom from re-operation will be analyzed.

#### Adverse Events

An adverse event (AE) is any unwanted clinical occurrence in a study participant, irrespective of whether it is associated with the study intervention or not. Pre-existing conditions are not considered adverse events unless there is a change in their nature, severity, or degree.

The incidence of serious adverse effects over the course of the trial will be compared between the two treatment groups. Specialists in charge of clinical control at each center will adjudicate all serious and protocol-defined AE. The safety endpoints will be recorded as the frequency of each adverse event, the rate of adverse events per patient/year and the time to each event. Additionally, the number of patients experiencing each type of serious adverse event will be recorded. Safety data will be collected during patient follow-up and the incidence of each event type will be calculated with 95% confidence intervals.

### Serious Adverse Event

According to Federal and Drug Administration guidelines, a serious adverse event is any experience that results in death, is life-threatening, results in significant or prolonged disability, requires or prolongs hospitalisation, results in a congenital anomaly/birth defect, or, in the opinion of the investigators, represents other significant risks or potentially serious injury to subjects or others. Major medical events that are not fatal, life-threatening, or requiring hospitalisation may be considered serious adverse events if, in the reasonable medical judgment of a clinician, they may endanger the patient and may require medical or surgical intervention to prevent one of the outcomes listed in this definition. Examples of medical events that may require medical or surgical treatment are: bronchospasm requiring intensive care in an emergency department or at home, blood dyscrasias, or seizures that do not require inpatient hospitalisation.

### Unexpected major adverse events

An unexpected major adverse event refers to a serious adverse event that is not defined in the protocol or documented in the patient consent form. Unscheduled notification is mandatory for unexpected serious adverse events.

During follow-up, the study will document any adverse events. The investigator will evaluate the relationship between the adverse event and the procedure performed. Causality is defined in **table 5**.

**Table 5.** Canadian Cardiovascular Society Classification (CCSC)†.

| Clinical Findings | Characteristics | Grade |
| --- | --- | --- |
| No limitation of ordinary activity | Normal physical activity (such as walking or climbing stairs) does not trigger angina. Angina can occur with intense, rapid or prolonged exercise at work or during leisure activities. | I |
| Slight limitation of ordinary activity | Angina may occur with<br>• walking or climbing stairs rapidly.<br>• walking uphill.<br>• walking or stair climbing after meals or in the cold in the wind or under emotional stress, or only during the few hours after awakening.<br>• walking more than 2 blocks on the level at a normal pace and in normal conditions .<br>• climbing more than 1 flight of ordinary stairs at a normal pace and in normal conditions. | II |
| Marked limitation of ordinary physical activity | Angina may occur after<br>• walking 1-2 blocks on level ground.<br>or<br>• climbing 1 flight of stairs at a normal pace. | III |
| Unable to carry on any physical activity without discomfort | Angina may occur when the patient is resting | IV |

†The Canadian Cardiovascular Society Classification of Angina Pectoris divides patients with angina symptoms into groups based on the severity of their symptoms. The classification uses the degree of restriction of day-to-day functions and the type of physical activity that triggers the angina episode. CCSC; Canadian Cardiovascular Society Classification

### Reporting of Serious Adverse Events

All investigators conducting clinical studies supported by the NHLBI must report both expected (protocol-defined) and unexpected serious adverse events. All serious adverse events must be reported directly to the clinical center’s IRB and the Data Coordinating Center (DCC) within 10 working days of knowledge of the event, or as dictated by the specific IRB policy, whichever is sooner. All deaths and unexpected serious adverse events must be reported to the DCC and the clinical center’s IRB within 24 hours of knowledge of the event, or as dictated by the specific Institutional Review Board (IRB) policy, whichever is sooner.

#### Specific Adverse Event Definitions

Myocardial infarction (MI) is diagnosed when there is clinical sign of myocardial necrosis consistent with myocardial ischaemia. [23] The diagnosis can be made if any one of the following criteria is met:

##### Myocardial Infarction

Evidence of an increase and/or decrease in cardiac biomarkers (ideally troponin) with at least one value above the 99th percentile of the upper limit of reference (URL), together with evidence of myocardial ischaemia with at least one of the following:

`o` ECG changes indicating new ischemia, such as new ST-T changes or new left bundle branch block (LBBB).
`o` Ischemia signs
`o` Pathological Q waves have developed in the ECG.
`o` Imaging findings of new loss of viable myocardium or new regional wall motion abnormalities.

##### Peri-CABG Myocardial Infarction

For patients undergoing CABG with normal baseline troponin values, elevations of cardiac biomarkers above the 99th percentile URL indicate peri-procedural myocardial necrosis. Conventionally, biomarker elevations greater than 5 times the 99th percentile URL, in addition to new Q waves or new LBBB, new graft or native coronary artery occlusion documented by angiography, or imaging findings of new loss of viable myocardium, have been considered to diagnose CABG-related MI.

##### PCI Myocardial Infarction

For patients undergoing PCI with normal baseline troponin values, elevations of cardiac biomarkers above the 99th percentile URL indicate peri-procedural myocardial necrosis. Elevations in biomarkers > 3 x 99th percentile URL have been conventionally interpreted as defining PCI-related MI. A subset associated with documented stent thrombosis is recognised.

Sudden unexplained cardiac mortality associated with cardiac arrest, often with symptoms consistent with myocardial ischaemia, and associated with suspected new ST elevation or new LBBB and/or evidence of fresh thrombus by coronary angiography and/or autopsy, where death precedes the collection of blood samples or the expected appearance of cardiac biomarkers in the blood, is classified as mortality due to MI.

#### Cardiac Arrhythmias

Any recorded arrhythmia that causes clinical impairment (e.g. haemodynamic impairment, oliguria, pre-syncope or syncope), necessitates hospitalisation or a physician visit, or occurs during hospitalisation. Cardiac arrhythmias are classified into two categories:

- Persistent ventricular arrhythmia necessitating defibrillation or cardioversion
- Persistent supraventricular arrhythmia necessitating drug treatment or cardioversion

#### Right-sided heart insufficiency

Persistent right ventricular dysfunction is indicated by a central venous pressure (CVP) greater than 18 mmHg and a cardiac index less than 2.0 L/min/m2, in the absence of elevated left atrial/pulmonary capillary wedge pressure (> 18 mmHg), tamponade, ventricular arrhythmias or pneumothorax. Treatment may require RVAD implantation, inhaled nitric oxide or inotropic therapy for more than 7 days.

#### Major Infection

A new episode of clinical infection with pain, fever, drainage and/or leukocytosis in the setting of antimicrobial treatment (non-prophylactic). There should be a positive culture from the infected site or organ, unless there is strong clinical evidence that treatment is required despite negative cultures. The following are the general categories of infection:

##### Localized Infection

The infection is limited to a specific organ or region, such as mediastinitis, and there is no evidence of systemic involvement (as defined by sepsis). This is determined through standard clinical methods and may be associated with evidence of bacterial, viral, fungal, or protozoal infection, or may require empirical treatment.

##### Endocarditis

The following clinical signs, symptoms, and laboratory tests are compatible with endocarditis: fever of 38. 0°C or higher, positive blood cultures, new regurgitant murmurs or heart failure, the presence of embolic phenomena (such as focal neurologic impairment, glomerulonephritis, renal and splenic infarcts, and septic pulmonary infarcts), and peripheral cutaneous or mucocutaneous lesions (such as petechiae, conjunctival or splinter hemorrhages, Janeway lesions, Osler’s nodes, and Roth spots). If echocardiography reveals a new intra-cardiac vegetation, along with or without other signs and symptoms, this should be considered sufficient evidence to support the diagnosis of endocarditis.

Transesophageal echocardiography (TEE) is the preferred diagnostic modality for prosthetic valve endocarditis.

##### Sepsis

There is sign of systemic involvement by infection as evidenced by positive blood cultures and/or hypotension.

#### Neurologic Dysfunction

Any new neurological deficit, whether transient or permanent, focal or global, identified by a standard neurological examination (performed by a neurologist or other qualified physician and documented by appropriate diagnostic tests and consultation notes). The investigating physician distinguishes between transient ischaemic attack (TIA) and stroke, TIA being fully reversible within 24 hours without evidence of infarction and stroke lasting more than 24 hours or less than 24 hours with evidence of infarction. To document the presence and severity of neurological deficits, the Modified Rankin Scale and the NIH Stroke Scale must be administered at the time of the event (within 72 hours following the event) and 90 days following the event.

Subcategories must be assigned to each neurological event as:

- A transient ischaemic attack (TIA) refers to an acute event that is completely reversed within 24 hours and does not show imaging evidence of infarction.
- Ischemic or Hemorrhagic Stroke, also known as Cerebrovascular Accident, is considered to be an event lasting more than 24 hours or less than 24 hours and accompanied by infarction on imaging. If an ischemic stroke undergoes hemorrhagic conversion, it should still be classified as ischemic.
- Toxic Metabolic Encephalopathy refers to a disturbance of brain function resulting from abnormal systemic metabolism or exogenous agents that alters awareness and/or consciousness, in which there is a non-focal neurological examination and a negative brain scan.
- Other

#### Renal Events

The classification of renal function will be based on the estimated glomerular filtration rate (eGFR), which will be calculated using both the Chronic Kidney Disease Epidemiology Collaboration (CKD-EPI) equation [24] and the European Kidney Function Consortium (EKFC) equation [25]. The severity of renal failure will be classified into different stages, as listed in **Supplementary Table 2.**

Two types of kidney related events will be recognised:

##### Renal Dysfunction

Abnormal kidney function is defined as a rise in serum creatinine (Cr) of more than 100% from baseline, and a Cr level greater than 2.0.

##### Renal Failure

New requirement for hemodialysis related to renal dysfunction. This definition excludes aquapheresis for volume removal alone.

#### Hepatic Dysfunction

If any two of the following liver test results (total bilirubin, aspartate aminotransferase/AST and alanine aminotranferease/ALT) are three times higher than the upper limit of normal for the hospital, or if liver dysfunction is the main cause of death.

#### Respiratory insufficiency

Respiratory function impairment that requires re-intubation, tracheostomy, or the inability to discontinue ventilatory support within 48 hours after surgery. This does not include intubation for re-operation or temporary intubation for diagnostic or treatment purposes.

#### Bleeding

A bleeding incident is characterised by any of the following

- Death due to hemorrhage can occur when more than 10 units of red blood cells are transfused within the first 24 hours following surgery.
- Re-operation may be necessary in cases of hemorrhage or tamponade.

NOTE: A hemorrhagic stroke is classified as a neurological event rather than a distinct bleeding event

#### Pericardial Fluid Trapping

Pericardial effusion is the collection of fluid or thrombus in the pericardial space requiring surgical intervention or percutaneous catheter drainage. This event can be further classified into two categories: those with clinical signs of tamponade (such as increased central venous pressure and decreased cardiac output) and those without signs of tamponade.

#### Arterial Non-CNS Thromboembolism

Confirmation of an acute systemic arterial perfusion deficit in any non-cerebrovascular organ system due to thromboembolism can be made by one or more of the following:

- Standard clinical and laboratory testing o Operative findings
- Autopsy findings

This definition excludes neurological events.

#### Wound Dehiscence

Surgical incision disruption, not caused by infection, that needs to be surgically repaired.

#### Venous Thromboembolic Event

Clinical and laboratory evidence of a venous thromboembolic event, such as deep vein thrombosis or pulmonary embolism, can be detected using standard tests.

#### Other

An occurrence that results in a clinically significant alteration in the patient’s state of health, or any event that is life-threatening, results in death, leads to permanent disability, necessitates hospitalisation or prolongs an existing hospital course.

### 2.4. Statistical methods

Continuous variables will be reported as mean and standard deviation or median and interquartile range as needed. Dichotomous and nominal variables will be reported as counts and percentages. Univariate analysis will be performed using the Mann–Whitney U test, Student’s t-test, Kruskal Wallis test, Wilcoxon test, Fisher exact test, Chi-square test and Kaplan-Meier test. Missing data will not be replaced. Multivariable analyses will be performed using logistic and the Cox-proportional hazards method. Variables’s selection for logistic regression will be done through the Lasso-based variable selection methods. Significant differences between study groups will be adjusted by using propensity score weighting. A doubly robust method (a combination regression model with inverse probability treatment weighting (IPTW) by propensity score) will be used to estimate the causal effect of the exposure on the outcomes. For this purpose, a covariate balancing propensity score will be developed to minimize the differences between groups.

A sample size of 630 patients, 384 for surgical mitral valve repair and 246 for transcatheter mitral valve repair, was calculated as sufficient to detect a significant difference in the rate of reverse left ventricular remodelling [80% power and α = 0.05 (two-tailed)]. A P value less than 0.05 is considered statistically significant. Statistical analyses were performed using R software (R Foundation for Statistical Computing, Vienna, Austria).

## 3. Ethics and dissemination

### Implications for treating patients undergoing Transcatheter versus Standard Surgical Mitral Valve Operation for Secondary Mitral Regurgitation

Analysis of the data from this registry will provide contemporary results for a large number of SMR patients with a long follow-up period. The risk of bias related to institutional volume and surgical experience is expected to be reduced by the multicenter nature of this registry. All participating centers must have an annual rate of at least 25 procedures for SMR and a mitral surgery programme that allows for proper follow-up and management of any late mitral events after replacement surgery or primary repair of the SMR. We expect to collect data that will provide insights into the impact of different surgical approaches on standard mitral valve surgery and TEER. This will help evaluate ventricular remodelling in the long-term follow-up. In addition, this report highlights the long-term mortality of patients undergoing TEER and provides conclusive results on the true worth of different standard mitral valve surgical strategies.

In detail, the results of the analysis comparing S-SMVp involving three different procedures and TMVp for SMR treatment can be summarized as follows, thanks to the multicentre nature of the registry:

- Specifically, it asks which procedure leads to the best outcome for LV remodelling and improved LVEF over a 10-year period.
- Whether death from all causes differs between cohorts at 10 years after S-SMVp or TMVp
- Whether patients with improved LVEF also have improved HF symptoms
- Which of the two procedures (S-SMVp and TMVp) achieves an immediate reduction of MR to moderate or lower levels and what percentage of patients can benefit from this result.
- The two groups are compared regarding the percentage of patients who experienced residual MR progression during follow-up. Whether patients with MR progression present with more severe heart failure symptoms.

### The lack of reliable data in current guidelines was the driving motivation for this study

Few observational studies have compared S-SMVp and TMVp for the treatment of SMR. [13, 16–18, 26,27] In a subgroup of the randomized trial Endovascular Valve Edge-to-Edge Repair Study (EVEREST), S-SMVp using RMA and TEER were also compared. [26] Among the subgroup of patients with SMR in EVERESTI trial (n = 56), there was no significant difference between TEER and RMA in terms of the composite endpoint of death, MV surgery or reoperation rate, and 3+ or 4+ SMR at 5 years (28.6% vs 40.5%; P = .43). [26] However, these findings were obtained from patients who underwent various surgical techniques, including repair and replacement.

Kortlandt et al. [27] compared 365 SMR patients treated with TEER with 95 patients who underwent MV surgery. They found comparable adjusted survival up to 3 years (HR, 0.86; 95% CI, 0.54-1.38; P = 0.541).

Two studies [17,18] compared S-SMVp using RMA with TEER in unmatched SMR patients. In a retrospective study of 76 patients treated with RMA and 95 patients with TEER, RMA resulted in a better reduction of MR and comparable adjusted survival at 6 months. [17] Similarly, in another study, 65 patients were treated with RMA and 55 with TEER. RMA was associated with a greater and more durable reduction of MR and comparable crude survival outcome at a median follow-up of 4 years. [18]

Recently, Okuno and colleagues [16] compared surgical repair with RMA to TEER for SMR and reported 2-year outcomes. The study highlights inconsistencies in the 2020 American Heart Association/American College of Cardiology (AHA/ACC) guidelines for the indication of TEER in SMR, which was class IIb with level of evidence B-R.2. [3] In this report of 202 patients, published immediately after the release of the new AHA/ACC guidelines, the investigators evaluated surgical versus transcatheter repair for SMR in a propensity-matched population. After a 2-year follow-up, the authors observed no significant difference in survival (P=.909), but RMA with coronary revascularisation was superior to TEER in reducing MR, improving ventricular ejection fraction, and reducing New York Heart Association functional class III or IV. (1)Moreover, Okuno and colleagues1 found that RMA was better than TEER after 2 years as a secondary endpoint. The Cardiothoracic Surgical Trials Network trial (CTSN) and Papillary Muscle Approximation (PMA) RCT trial provided evidence that RMA had higher MR recurrence rates at 2 and 5 years of follow-up, with rates of 58.8% and 55.9%, respectively. [10,28]

Currently, there is limited data on the effectiveness and safety of RMA in treating patients with HF and SMR. Existing studies are small observational studies that suggest improvement in LV function and functional status (29-31). However, the line confirming the suitability for RMA should include seventy-four patients from the CTSN trial. The cohort has preoperative left ventricular end-systolic diameter and reduced apical leaflet tethering. At the 2-year follow-up, patients with severe ischemic mitral regurgitation who did not experience persistent or recurrent MR after RMA had significantly smaller left ventricles (43±26 mL/m2) compared to those who did (63±27 mL/m2). Additionally, their left ventricular end-systolic volume was significantly lower than that of patients who underwent mitral valve replacement (61±39 mL/m2). [28]

LV remodeling is a predictor of a worse prognosis in ischaemic heart disease and is reversible with recovery of viable myocardium. [10,32] In the CTSN study, 75% of patients underwent CABG at the same time, precluding the potential for improved regional wall motion in 25% of patients. [28] Previous studies have shown that RMA+SPM-r is superior to RMA alone for both ischemic and nonischemic cardiomyopathy. [10,11,32,33] In the PMA RCT, 96 patients with severe chronic ischaemic mitral regurgitation who underwent complete surgical myocardial revascularisation were assigned to receive either isolated RMA or PMA plus RMA, followed for 5 years. At the 5-year follow-up, there was an improvement in the left ventricular end-diastolic diameter (5.8 ± 4.1 mm and - 0.2 ± 2.3 mm, respectively; P < .001). The benefit achieved immediately postoperatively was maintained, with freedom from major adverse cardiac events (P = .004) [10]

The COAPT study found no improvement in left ventricular (LV) remodeling with the use of transcatheter edge-to-edge repair (TEER). The left ventricular end-diastolic volume (LVEDV) was 194.4±69.2 vs. 192.2±76.5 mL [14,34]. However, TEER was found to provide better outcomes in terms of symptom relief and quality-of-life measures when compared to guideline-directed medical therapy (GDMT) alone. In a study of 58 patients who had previously only received GDMT and then underwent TEER procedure, it was observed that the composite rate of death or hospitalization for HF was lower in this group compared to those who only received GDMT (P=0.006). [34]Additionally, TEER recipients reported improvements in mitral regurgitation (MR) severity and functional capacity after 3 and 5 years. [15,34]

All five 2020 AHA/ACC recommendations were rated LOE B-R or B-NR. None of these recommendations are based on reports with 5-year follow-ups, indicating moderate study quality. [3,13] The available literature lacks randomized controlled trials with large numbers of enrolled patients, including candidates for TEER, mitral valve replacement, or mitral valve repair with or without subvalvular procedures. The only references to TEER-based RCTs with 3-year outcomes are the COAPT trial [13,34] and the analysis of the new pathophysiological framework of the pathomechanism for SMR [13,35] in the ACC/AHA guidelines. Currently, there is no solid evidence for subvalvular repair. [3,13]

## Data Availability

All data produced in the present study are available upon reasonable request to the authors

https://ccn.fr/

## Acronyms

AE: adverse event
CABG: coronary artery bypass surgery
CCSC: Canadian Cardiovascular Society Classification
CHF: chronic heart failure
COAPT: cardiovascular Outcomes Assessment of the MitraClip Percutaneous Therapy
GDMT: Guideline-directed medical therapy
HF: heart failure
ICU: intensive care unit
LCP: Leaflet and chordal procedures
LV: left ventricle
LVEDP: Left ventricular end-diastolic pressure
LVEF: Left Ventricular Ejection Fraction
MACE: Major Adverse Cardiac Events
MCG: Medical College of Georgia
MI: Myocardial infarction
MV: mitral valve
NYHA: New York Heart Association
PCI: percutaneous coronary intervention
PCWP: Pulmonary capillary wedge pressure
RCT: randomized clinical trials
RER: respiratory exchange ratio
RMA: Restrictive mitral annuloplasty
RPE: rate of perceived exertion
SMR: Secondary mitral regurgitation
S-SMVp: standard surgical mitral valve procedure
TEER: transcatheter edge-to-edge mitral valve repair (TEER)
TEERMISO: transcatheter Versus Standard Surgical Mitral Valve Operation for Secondary Mitral Regurgitation
TMVp: transcatheter mitral valve procedure

## Authors’ contributions

state how each author was involved in writing the protocol.

