## Supplementary_material for "A study protocol for an international registry observational study evaluating clinical outcomes of transcatheter versus standard surgical mitral valve operation for secondary mitral regurgitation: the TEERMISO study"

**S1- Clinical Centers**

The study will take place in all five clinical centers participating in the TEERMISO protocol trial. Each clinical centre will be responsible for promptly obtaining IRB approval of the protocol and informed consent (including amendments), enrolling patients, accurately collecting data and entering it into the electronic data capture system (Microsoft Access datasheet; Redmond, Washington, USA ). The coordinating centre will be located in France at the Centre Cardiologique du Nord, which will be the NCT from Clinical Trial Gov. Com (**ID: NCT05090540)** and the IRB (**ID : 202201143)** as the primary trial centre.

- **Profile of the investigator, skills and expertise**

All surgeons, cardiologists, coordinators, and other investigators involved in the study are required to complete the Investigator Profile form. The form should include their hospital affiliation, address, telephone number, fax number, and email address. The surgeon, cardiologist, and coordinator are required to email their Conflict of Interest Statement and Financial Disclosure Certification. A staff qualified in administrative health procedures will be authorised to assist the specialists in carrying out the study.

The clinical investigators for this trial are cardiothoracic surgeons and interventional cardiologists who specialise in surgical repair of the mitral valve and transcatheter valve procedures, and who are experienced in the care of patients with ischaemic and valvular heart disease. Specifically, to be eligible as a participating surgeon, they must have performed at least 15 mitral valve procedures per year, averaged over a 2-year period. Interventional cardiologists must have undergone specific training in the use of transcatheter therapies on new platforms for the treatment of structural heart valve disease.

- **Conflict of Interest and Financial Disclosure Statement**

This statement confirms that all investigators involved in this study have no conflicts of interest with any institution that may affect their participation. Each investigator must complete this statement and submit a financial disclosure agreement.

- **Patient privacy**

Patient records will be kept confidential in accordance with HIPAA guidelines. (<https://www.hhs.gov/hipaa/index.html>). Source documentation may be reviewed by Study Investigators, site Institutional Review Boards (IRBs), or single centre authorization by local ethics committee. However, all unique patient and hospital identifiers will be removed. Aggregate data from this study may be published in accordance with the publication policy documented in the trial agreements. However, no data with patient identifiers will be published. After the data has been included, the statistician in charge of the analysis will analyse both the original ACCESS datasheet and the extracted Excel (Redmond, Washington, USA ). He or she will then contact the principal investigator of each centre involved in the study to ensure that privacy is respected, and a single database will be used for the final analysis.

- **S2-** **Mechanical intervention performed in group patient**

Patients will be enrolled in either (a) Standard Mitral Valve Procedure (S-SMVp), which involves mitral valve repair (RMA or RMA plus SVR) or MVR, or (b) Trancatheter Mitral Valve Procedure (TMVp), which involves TEER.

a) ***Standard Mitral Valve Procedure***

After induction of anaesthesia, all patients will undergo transesophageal echocardiography (TEE) and pulmonary artery catheterisation according to centre preference and cardiothoracic anaesthesia protocol. Central hemodynamics will be recorded, and TEE will be performed under loading conditions as close as possible to the patient's baseline. Intra-operative TEEs will be performed according to standardized procedures (refer to echo protocol appendix ), which will also be provided to the site and affixed to the operating room echo machine.

To prevent anaesthetic effects, maintain blood pressure and pulmonary artery pressures using phenylephrine or 2,6-diisopropylphenol and volume during TEE evaluations. Confirmation of tethering of the anterior and posterior leaflets by the posterior papillary muscle and adjacent left ventricle was observed in patients who underwent MV isolated RMA or combined RMA plus SVR annuloplasty.

Depending on institutional routines, procedures are performed with either full or partial sternotomy and cardiopulmonary bypass. Double venous cannulation may be performed either by percutaneous or by direct venous access. Depending on the surgeon's preference, antegrade and/or retrograde cardioplegia will be used and coronary artery bypass grafting will be performed. The surgeon will evaluate the adequacy of the revascularization.

To approach the mitral valve, one can use either the left atrium via Waterston's groove or a biatrial approach. It is important to inspect the mitral valve to confirm the absence of any organic pathology, such as myxomatous, rheumatic, or endocarditic changes.

- **Coronary Artery Bypass Grafting (CABG)**

Standard techniques will be used to perform coronary artery bypass grafting with two-stage venous cannulation. The LIMA is recommended for LAD grafts, but conduit selection and harvesting methods will not be prescribed. Technical details of bypass grafting will not be specified. The surgical investigator will perform complete revascularization based on their judgment.

- **Mitral valve repair using isolated RMA or combined RMA plus SVR**

Mitral valve exposure has been via a Sondergaard or Waterston groove or via a transseptal (biatrial) approach. A complete and rigid or semi-rigid mitral annuloplasty ring from any manufacturer was placed circumferentially in the mitral valve annulus using non-pledgeted 2-0 braided sutures. Usually, four to six sutures are placed in the anterior annulus and six to eight sutures in the posterior annulus. The annuloplasty ring size is determined based on the surface area of the anterior mitral leaflet, which is measured by determining the intertrigonal distance and anterior leaflet height. Typically, annuloplasty ring sizes ranging from 24 to 28 mm are used, which are 2 ring sizes smaller than the measurement. For males, this will be an average of 28mm, and for females, it will be an average of 26mm. The ring is sutured with appropriate spacing and then lowered into position before the sutures are tied. Finally, to check for any residual regurgitation, the ventricle is pressurised with saline.

If the surgeon deems it necessary due to severe tethering, an additional subvalvular procedure may be added based on their judgement and experience. In patients requiring SVR, mitral geometric abnormalities were identified by changes in three metrics: anteroposterior annular dilatation, tenting area and interpapillary muscle distance. Leaflet tethering caused by excessive traction resulting in lack of coaptation was found in all patients. To confirm the echocardiographic findings, the surgeon measured the inter-papillary muscle distance (IPMD) according to center experience and habit. It is important to note that papillary muscles identified anatomically as type I and II were approximated using a CV-4 Gore-Tex suture (W. L. Gore & Associates, Flagstaff, Ariz). The stitch was positioned on the head of each PM **(supplementary Figure 2).** For type III, IV, or V PMs, a 4-mm Gore-Tex tube was used to encircle the bodies of the posteromedial and anterolateral papillary muscles. The two separate heads were then reinforced together, as shown in Figure 1S. This approach allowed for the simultaneous approximation of both posteromedial papillary muscles, which reduced mitral valve tenting. [1-7]

The evaluation of the precise anatomical location of chordae tendinea was crucial in improving regurgitation correction. Special attention was given to this arrangement. The chordae tendineae from the posterior papillary muscle (PPM) are typically inserted on the P2 and P3 cusps of the posterior leaflet, whereas those from the APM are destined for the anterior leaflet, contributing primarily to the development of the 'seagull sign' and the associated tenting. [1-7] **Supplementary Figure 2**

There are two circumstances in which a mitral valve repair may be crossed over to a mitral valve replacement: (1) if the valve is deemed unrepairable on direct assessment during the operation, or (2) if the TEE shows more than mild residual MR. The degree of residual MR is measured on the immediate post-cardiopulmonary bypass (CPB) TEE. [8]

The closure will involve the placement of standard epicardial atrial and ventricular pacing wires based on the surgeon's preferences. Mediastinal and pleural drains will also be inserted and removed according to institutional routines. Perioperative hemodynamic support may be required and will be determined by the surgeon.

- **Mitral valve Replacement**

The Khonsary procedure [9-11] is the most commonly used approach for complete chordal-sparing mitral valve replacement. To expose the mitral valve, the Sondergaard or Waterston groove or a transseptal approach involving the biatrial way can be used with partitioning of the anterior mitral leaflet. Pledgetted 2-0 braided sutures were placed to include the mitral annulus and the partitioned sections of the anterior leaflet, resulting in plication of the chords to the annulus. For the posterior annulus, incorporate both the annulus and the posterior leaflet with sutures, imbricating any redundant tissue. The sizing process involves using the manufacturer's sizers. If redundant anterior leaflet tissue is present, to avoid LV outflow tract obstruction or interference with the mechanical valve mechanism, excise a central elliptical portion of the anterior leaflet, leaving 5-10 mm of leaflet free edge attached to the primary chordae tendineae. The mitral annulus size is measured using standard valve sizers. The prosthesis is then repositioned and sutured. The final step is confirmation of free movement of the prosthesis leaflets. If a mechanical prosthesis is used, perform valve rotation as needed. The choice between a mechanical or bioprosthesis is left to the surgeon's discretion, but a mitral bioprosthesis is usually sufficient for most patients.

- **Indications for surgical tricuspid valve annuloplasty**

In cases of severe tricuspid insufficiency, defined as follows, tricuspid repair will be performed concomitantly with the mitral procedure.

The preoperative awake transthoracic echocardiogram (TTE) shows severe tricuspid regurgitation caused by tricuspid annular dilatation. The leaflets and subvalvular apparatus are structurally uninvolved. Filling pressures are normal for age (systolic systemic blood pressure > 90 mmHg, pulmonary diastolic pressure > 10 mmHg). Severe TR is defined by a maximum jet area > 10 cm or > 66% of the RA area. In the presence of pulmonary hypertension with mean pulmonary artery pressure >50 mmHg, tricuspid repair is a class IIa recommendation for moderately functional TR (maximum jet area 6-10 cm or 33-66% of RA area) secondary to a left heart lesion at the time of mitral valve surgery.

Repair of the tricuspid valve for moderate functional insufficiency (maximum jet area of 6 to 10 cm or 33 to 66% of the right atrial area) secondary to left-sided pathology during mitral valve surgery is a grade IIa recommendation. The decision to perform a tricuspid procedure in these patients is left to the surgeon's personal judgement and may be addressed as part of a sub-study or separate protocol after further consideration by the heart team at each centre for shared decision making. [12]

- **Tricuspid annuloplasty surgical procedure**

A tricuspid anuloplasty system, such as the Carpentier-Edwards annuloplasty ring or the Edwards MC3, should be used. Using commercially available gauges, the size of the prosthetic ring should be determined based on the distance between the antero-septal and postero-septal commissures and the anterior leaflet surface area. Typically, a 30-32 ring is preferred for men and a 28-30 ring is preferred for women. The fibrous tricuspid annulus is sutured with eight to ten U stitches, each 1 cm wide, placed 2 mm from the leaflet hinge, except in the area of the bundle of His located in front of the first half of the septal leaflet. Plication of the annulus is achieved with smaller distances applied to the ring in front of the anterior and posterior leaflets. After dropping the ring and tying the sutures, the appropriateness of the repair is verified by the saline infusion check. If the repair is deemed unsatisfactory or the valve is deemed unrepairable by the surgeon, replacement of the valve may be necessary.

b) ***Trancatheter Mitral Valve Procedure***

Transcatheter mitral valve repair (TMVr) is achieved using the MitraClip Procedure. Warfarin should be discontinued for at least three [13,14] days prior to the scheduled MitraClip procedure to achieve an International Normalised Ratio (INR) ≤ 1.7. Similarly, dabigatran or Factor Xa inhibitors should be discontinued for a sufficient period to ensure restoration of normal coagulation. Heparin may be used during this period at the treating physician's discretion. If heparin is used, subcutaneous low molecular heparin (LMWH) should be stopped 12 hours before MitraClip and intravenous unfractionated heparin (UFH) should be stopped 4 hours before MitraClip. A loading dose of clopidogrel at a dose determined by the investigator should be started either immediately before or after the procedure. [13,14]

The COAPT and MITRA FR protocols require an additional transesophageal echocardiogram (TEE) within 3 days prior to the procedure to rule out the presence of intra-cardiac masses, thrombi, or vegetations. The echocardiogram can be performed just before starting the TEER procedure. If an intra-cardiac mass, thrombus, vegetation, or any other change in echocardiographic eligibility is detected on the TEE performed immediately prior to the mechanical intervention, the subject will not receive the device implantation. In case a blood clot is found, the patient may receive treatment to dissolve it. If successful, the patient should be reevaluated promptly to determine if they are a candidate for the TEER procedure. If a patient has a MitraClip device implanted and undergoes a subsequent procedure involving the device, the follow-up schedule will be based on the time of the initial implantation. After crossing the transseptal, administer intravenous heparin or alternative anticoagulation therapy according to standard hospital practice. Maintain an activated clotting time of over 250 seconds throughout the procedure. [13,14]

The steerable guidewire catheter (Guide) is introduced into the femoral vein and advanced across the transseptal puncture site. Fluoroscopic and echocardiographic guidance will be used during the procedure for visualisation of the devices, vasculature and cardiac anatomy. The MitraClip Delivery System is placed into the guide and positioned properly over the MV . The MitraClip Delivery Catheter forwards until the MitraClip device protrudes from the tip of the guide into the left atrium. The catheter tip should be manipulated using the control knobs on the handles until the MitraClip device is properly positioned perpendicular to the coaptation line of the MV. The MitraClip device is advanced across the MV into the ventricle LV and then retracted to grasp the leaflets. Two-dimensional and 3-dimensional echocardiography, along with color flow Doppler, are utilised to assess the presence of a double orifice, leaflet insertion, MitraClip device positioning, and residual MR. If the MitraClip device is not positioned correctly or if MR has not been sufficiently reduced, additional grasping attempts may be made. The MitraClip device may also be inverted in the left atrium if necessary. Once placement is successfully achieved, the MitraClip device is closed and deployed from the Delivery Catheter. The catheters are then retrieved from the patient. If the placement of one MitraClip device does not sufficiently reduce MR to an acceptable level, a second device may be used to further reduce it. The investigator must ensure that there is enough space in the mitral valve orifice to accommodate the second device without causing stenosis. This trial permits the implantation of a maximum of three MitraClip devices per subject. [13,14]

Following the MitraClip procedure, anticoagulant treatment, e.g. heparin, is discontinued and ACT is monitored according to hospital protocols and international guidelines. [15,16] According to usual hospital practice, vascular sheaths will be removed. Subjects will receive standard care following cardiac catheterization, as deemed appropriate by the investigator. For the first month after the MitraClip procedure or longer, as deemed appropriate by the investigator, subjects should be instructed to limit vigorous physical exercise.

Provide additional intravenous doses of antibiotics at approximately 6 and 12 hours (or according to institutional guidelines) after the procedure is considered to be finished. It is important to inform all patients receiving the MitraClip device about the need for endocarditis prophylaxis in accordance with the ACC/AHA 2020 update guidelines and [2023 ESC Guidelines for the management of endocarditis.](https://pubmed.ncbi.nlm.nih.gov/37622656/) [15,17] The subject may receive a prescription for endocarditis prophylaxis upon discharge. After the TEER is performed, anticoagulation therapy is prescribed. If a loading dose of clopidogrel was not administered prior to the MitraClip procedure, it should be given as soon as possible after the procedure. Anticoagulation after TEER procedure is as follows: 1) Recommence warfarin, dabigatran or factor Xa inhibitor at the same dosage as before the MitraClip procedure, or as directed or 2) Initiate a daily dose of 81-325 mg of acetylsalicylic acid (ASA) for a period of 6 months or longer, as determined by the Investigator and if deemed necessary. [15,16]

***Other therapeutics***

- **GDMT**

Patients should receive standard medical treatment for their coronary artery disease, mitral valve regurgitation, and any other coexisting conditions as per current medical guidelines. This may include beta-blocker therapy, angiotensin converting enzyme (ACE) inhibitors or ARBs, platelet aggregation inhibitors, statin therapy, aldosterone antagonists, implantable cardiac defibrillators and cardiac resynchronisation therapy if clinically appropriate and well-tolerated. The guidelines for medical management will be updated by a clinical management committee by each heart team [15,16]

- **Managing Postoperative Atrial Fibrillation (AF)**

Post-operative AF will be assessed by the following criteria. To control the ventricular response rate in postoperative AF , beta-blockers are recommended as the first-line therapy for rate control. If beta-blockers are contraindicated, nondihydropyridine calcium channel blockers should be used as second-line agents. In patients with coronary artery disease who do not have congestive heart failure, converting to normal sinus rhythm can be achieved with Sotalol or class 1A antiarrhythmic drugs. For patients with depressed left ventricular function, amiodarone therapy is recommended. If postoperative AF atrial remains or is recurrent for more than 24 hours, anticoagulation with warfarin is advised for 4 to 8 weeks or at least 30 days after return of sinus rhythm. To reduce the incidence of AF , beta-blockers should be administered preoperatively or early postoperatively in patients without contraindications. In patients who cannot take beta-blockers, preoperative administration of amiodarone may reduce the incidence of postoperative atrial fibrillation. [13,14,18,19]

**Legend**

**Supplementary Figure 1** Study Design Schematic

**Supplementary Figure 2**. The figure shows a variety of surgical techniques for the management of the subvalvular apparatus (A to C). It includes the anatomy of the papillary muscles and corresponding surgical techniques. Type I is a single homogeneous entity, whereas type II has a groove with two peaks (blue arrow). Type III has fenestrations with muscular bridges. Type IV has complete separation in two adjacent heads, and type V has complete separation with two distant heads (purple arrow). PMA is treated with a 4-0 Gore-Tex suture for type 1 and 2 (A), and a 4-0 Gore-Tex prosthesis for type 3-5, which results in PM slinging (B and D). A 4-0 Gore-Tex suture is used to achieve PM repositioning. Attached to the trigone are the ALPM (green arrow) and the PMPM (red arrow). Two sutures were used to attach both tips of the PMPM to the posterior trigone to avoid the Segul sign in PMs type 2-5.

**Supplementary Table 1.** Inclusion and exclusion criteria for patients with SMR requiring mechanical intervention

**Supplementary Table 2** Criteria for defining postoperative acute kidney injury†

† Serum creatinine level changes during hospitalization
