## Supplementary material for "A study protocol for an international registry observational study evaluating clinical outcomes of transcatheter versus standard surgical mitral valve operation for secondary mitral regurgitation: the TEERMISO study": Table S1

**Supplementary Table 1. Inclusion/exclusion criteria for SMR requiring mechanical intervention**

**Inclusion Criteria**

- Age 18 years or older,
- Symptomatic secondary mitral regurgitation (3+ or 4+ by i echocardiographic laboratory assessment) due to cardiomyopathy of either ischaemic or non-ischaemic etiology.
- The subject has been adequately managed according to applicable guidelines, including for coronary artery disease, LV dysfunction and disease, LV dysfunction, mitral regurgitation and heart failure.
- Subjects with at least one hospitalisation for heart failure. BNP ≥300 pg/ml or a corrected NT-proBNP ≥1500 pg/ml.
- NYHA functional class II, III or ambulatory class IV.
- Each centre's heart team will decide whether to offer MV surgery or TEER.
- Left ventricular end-systolic dimension ≤70 mm
- Left ventricular ejection fraction ≥20% and ≤50%.
- The nature of the primary regurgitant jet is commissural or non-commissural. For the MitraClip group, the implanting investigator's opinion is that the MitraClip can successfully treat the jet (if a secondary jet is present, it must be considered clinically insignificant).
- Transseptal catheterisation and femoral vein access is possible with the MitraClip implanting investigator.
- The subject has given written informed consent and agrees to all the terms of the study and the follow-up visits after the procedure.

**Exclusion Criteria**

- Life expectancy is less than 12 months due to non-cardiac conditions.
- Excessive surgical risk (in the judgment of the surgical investigator)
- Patients who have undergone surgery on their mitral valve leaflet in the past, have a prosthetic mitral valve implanted, or have undergone a transcatheter mitral valve procedure prior to hospitalization.
- The subject must have undergone transcatheter aortic valve replacement (TAVR) within 30 days prior to hospitalization.
- Patients have had mitral valve surgery, prosthetic in the past, have a prosthetic mitral valve implanted, or have undergone a transcatheter mitral valve procedure before, please let us know.
- Surgery is required for tricuspid valve disease with the exception for tricuspid valve repair
- Aortic valve disease that requires surgery or transcatheter intervention.
- Status 1 heart transplant or previous orthotopic heart transplantation.
- Patients requiring emergency or urgent surgery for any reason.
- Preoperative Congenital heart disease (except PFO or ASD)
- In patients in whom transesophageal echocardiography (TEE) is contraindicated or high risk.
- Echocardiographic evidence of intracardiac mass, thrombus or vegetation.
- Patients with active endocarditis or active rheumatic heart disease or leaflets degenerated from rheumatic disease (i.e., noncompliant, perforated).
- Active infections requiring current antibiotic therapy.
- If patients are pregnant or plan to become pregnant within the next 12 months
- Chronic Obstructive Pulmonary Disease (COPD) and requires continuous home oxygen therapy or chronic outpatient oral steroid use.
- The subject must not have undergone coronary artery bypass grafting (CABG) within 30 days prior to hospitalization for TEER procedure
- Subject hospitalization is not allowed if the individual has had a cerebrovascular accident within the last 30 days.
- Severe symptomatic carotid stenosis, as determined by ultrasound, is greater than 70%.
- ACC/AHA Stage D heart failure.
- Chronic renal insufficiency defined by Cr ≥ 2.5 or chronic renal replacement therapy (chronic hemo- or peritoneal dialysis)
- If the estimated pulmonary artery systolic pressure (PASP) is greater than 70 mm Hg, as assessed by echocardiography or right heart catheterization, it is considered high unless active vasodilator therapy in the cath lab can reduce the pulmonary vascular resistance (PVR) to less than 3 Wood Units or between 3 and 4.5 Wood Units with v wave less than twice the mean of the pulmonary capillary wedge pressure.
- Structural heart diseases that can cause heart failure, excluding dilated cardiomyopathy of either ischemic or non-ischemic etiology. These diseases include hypertrophic cardiomyopathy, restrictive cardiomyopathy, and constrictive pericarditis.
- Infiltrative cardiomyopathies such as amyloidosis, hemochromatosis, and sarcoidosis.
- The patient is experiencing hemodynamic instability and requires inotropic support or mechanical heart assistance.
- The echocardiogram shows moderate or severe right ventricular dysfunction, indicating physical evidence of right-sided congestive heart failure.
- Implant of any Cardiac Resynchronization Therapy (CRT) or Cardiac Resynchronization Therapy with cardioverter-defibrillator (CRT-D) within the last 30 days prior hospitalization.
- The mitral valve orifice area should be assessed by a transthoracic echocardiogram (TTE) within 90 days prior to subject hospitalization and should be less than 4.0 cm2.
- Modified Rankin Scale ≥ 4 disability.
