## Supplementary material for "A study protocol for an international registry observational study evaluating clinical outcomes of transcatheter versus standard surgical mitral valve operation for secondary mitral regurgitation: the TEERMISO study": Table S2

| **Table 4** Criteria for defining postoperative acute kidney injury† [43,44] | |
| --- | --- |
| **Level** | **Creatinine in serum** |
| 1 | Increase of 1.5-1.9 times the baseline  or an increase of at least 0.3 mg/dL (26.5 micromol/L). |
| 2 | 2 to 2.9 times the baseline. |
| 3 | Increase to 3.0 times the baseline  or to 4 mg/dL (353.6 micromol/L) or  initiation of renal replacement therapy. |

† Serum creatinine level changes during hospitalization
