## Supplementary figures and images for "A study protocol for an international registry observational study evaluating clinical outcomes of transcatheter versus standard surgical mitral valve operation for secondary mitral regurgitation: the TEERMISO study"

### Figure S1

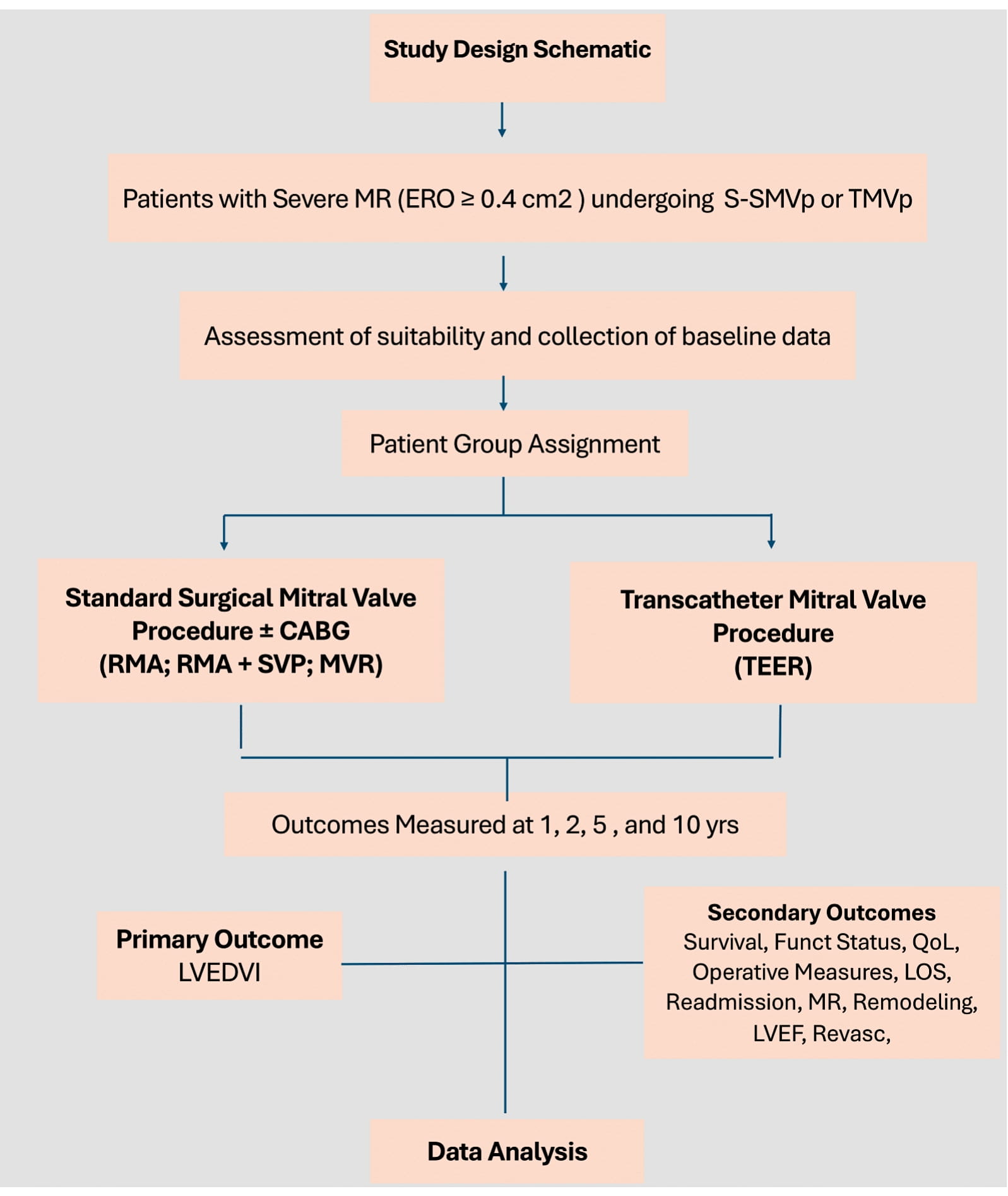

### Figure S2

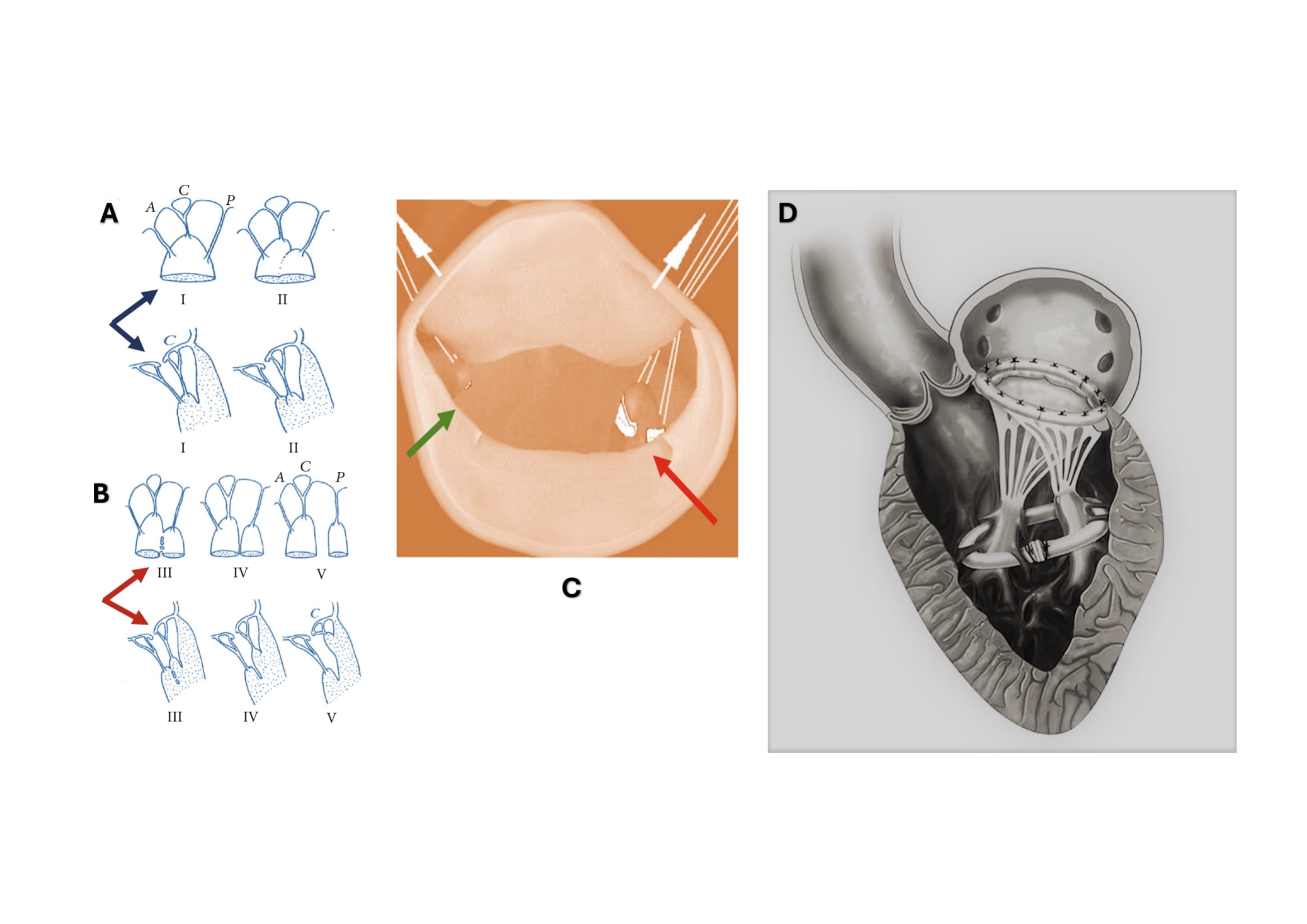
